# Outbreak of COVID-19 and Interventions in One of the Largest Jails in the United States — Cook County, IL, 2020

**DOI:** 10.1101/2020.07.12.20148494

**Authors:** Chad Zawitz, Sharon Welbel, Isaac Ghinai, Connie Mennella, Rebecca Levin, Usha Samala, Michelle Bryant Smith, Jane Gubser, Bridgette Jones, Kate Varela, Uzay Kirbiyik, Josh Rafinski, Anne Fitzgerald, Peter Orris, Alex Bahls, Stephanie R. Black, Alison M. Binder, Paige A. Armstrong

**Affiliations:** Cermak Health Services; Cook County Health; Chicago Department of Public Health; Centers for Disease Control and Prevention; Cook County Sheriff’s Office; University of Illinois at Chicago

## Abstract

**Background:** Correctional and detention facilities are disproportionately affected by COVID-19 due to shared space, contact between staff and detained persons, and movement within facilities of detained persons, many with pre-existing medical conditions. On March 18, 2020, Cook County Jail, one of the United States’ largest, identified its first suspected case of COVID-19 in a detained person.

**Methods:** This analysis includes SARS-CoV-2 cases confirmed by molecular detection among detained persons and Cook County Sheriff’s Office staff. We examined occurrence of symptomatic cases in each building and proportions of asymptomatic detained persons testing positive. We describe timing of interventions including social distancing, mask use, and expanded testing and show outbreak trajectory in the jail versus contemporaneous case counts in Chicago.

**Results:** During March 1–April 30, 907 symptomatic and asymptomatic cases of SARS-CoV-2 infection were detected among detained persons (n = 628) and staff (n = 279), with nine deaths. Symptomatic cases occurred in all housing divisions; in 9/13 buildings, staff cases occurred first. Among asymptomatic detained persons in quarantine, 23.6% tested positive. Visitation stopped March 15, programmatic activities were suspended March 23, cells were converted into single occupancy beginning March 26, and universal masking was implemented for staff (April 2) and detained persons (April 13). Cases at the jail declined while cases in Chicago increased.

**Conclusion:** Aggressive intervention strategies coupled with widespread diagnostic testing of detained and staff populations can limit introduction and mitigate transmission of SARS-CoV-2 infection in correctional and detention facilities.

## BACKGROUND

In correctional and detention facilities, shared physical space and interaction of detained persons and staff facilitate introduction and spread of viruses like SARS-CoV-2.^1^ Large COVID-19 outbreaks have been reported in congregate settings^2,3^, including correctional and detention facilities.^4^ Multiple interventions, including physical distancing and reducing introductions from the community via new detainees, staff, and visitors, are likely needed to effectively interrupt SARS-CoV-2 transmission, but can be difficult to implement.^5,6^ Many individuals incarcerated or detained in U.S. state and federal facilities are at elevated risk for severe COVID-19: they are more likely than the general population to be immunocompromised^7^ and approximately 50% have pre-existing medical conditions.^8^

Cook County Jail (CCJ) is one of the largest in the United States. On March 18, 2020, a person detained at CCJ reported influenza-like illness, including shortness of breath and fever, but tested negative for influenza. Cermak Health Services (CHS) medical staff suspected COVID-19, isolated the patient, and notified the Chicago Department of Public Health (CDPH). Although the patient did not meet COVID-19 testing criteria (no international travel or known exposure), CHS submitted diagnostic specimens to the Centers for Disease Control and Prevention (CDC). On March 28, a specimen tested positive for SARS-CoV-2 by real-time reverse transcriptase polymerase chain reaction (rRT-PCR).

We describe the subsequent outbreak of COVID-19 among detained persons and staff at CCJ and interventions to reduce transmission. CHS, the Cook County Sheriff’s Office (CCSO), Cook County Health (CCH), CDPH, and CDC partnered to investigate, identify, and interrupt transmission.

## METHODS

### STUDY POPULATION AND FACILITY CHARACTERISTICS

In 2019, approximately 59,000 people were admitted into custody at CCJ; the average daily number of detained persons was 5,800. During March 1–April 30, 2020, the population of detained persons declined from 5,579 to 4,054; average daily census was 4,884. On March 1, CCSO had 2,370 sworn personnel assigned to CCJ, representing the majority of staff who work at CCJ. During the outbreak, 270 sworn personnel were added to secure an expansion in CCJ’s geographic footprint.

CCJ houses detained persons in nine divisions in 13 buildings. Divisions are either open dormitories housing 40□ 48 individuals on average (though one dormitory can house >600 detained persons at full capacity), or units with double-occupancy cells and shared common spaces. Participation in programmatic activities (e.g., work assignments, school) and medical needs of individuals vary by division. Prior to March 1, 2020, CCJ utilized seven divisions; during March 1–April 30, two additional divisions were opened to achieve social distancing through single-cell occupancy and an alternating bunk model.

### DETECTION AND INTERVENTIONS

#### Case definition

COVID-19 cases were defined as persons with an epidemiologic link to CCJ and molecular evidence of SARS-CoV-2 infection during March 1–April 30, 2020.

#### Quarantine, medical isolation, and testing of symptomatic detained persons

Any detained person reporting symptoms consistent with COVID-19 was medically isolated in a single cell, assessed by medical staff, and tested for SARS-CoV-2 via rRT-PCR performed at Illinois Department of Public Health, QUEST diagnostics (Secaucus, New Jersey), or Stroger Hospital using the *m*2000 system (Abbott Laboratories, Illinois, USA). Beginning April 20, testing of newly detained persons was performed with the ID NOW™ COVID-19 assay (Abbott).

In the event a resident on a unit tested positive or ≥2 suspected cases were detected, the remaining individuals on the unit were placed under quarantine for ≥14 days; no individuals were moved onto or off the unit. Quarantined persons were assessed daily for symptoms; if any became symptomatic, they were medically isolated, and quarantine was extended an additional 14 days for the remainder of the unit.

#### Testing of asymptomatic detained persons

Testing was offered to asymptomatic detained persons in units placed under quarantine from March 25, beginning with individuals at increased risk of severe disease and extending to buildings with new and active cases.

#### Screening and testing of staff

Staff included only CCSO employees working on the CCJ campus. Staff were provided with a list of testing locations, but testing was optional. Staff were required to report symptoms, positive test results, or COVID-19 clinical diagnoses to CCSO; affected individuals were provided paid time off. Staff cases were cross-referenced with Illinois’ National Electronic Disease Surveillance System (I-NEDSS) to validate laboratory results.

### DATA ANALYSES

Epidemiologic curves by division were constructed using date of medical isolation as a proxy for date of symptom onset for detained persons, and by self-reported symptom onset date for staff. First positive specimen collection date was used if medical isolation date was not available.

The attack rate (AR) among detained persons at CCJ was calculated using bed assignments by day. Demographic and clinical data for fatal cases were abstracted from medical records, but were unavailable for other cases. Categorical data are presented as proportions; continuous data are presented as medians (interquartile range [IQR]). Data for all persons residing in the state of Illinois meeting the case definition for confirmed SARS-CoV-2 infection were extracted from I-NEDSS and included using the specimen collection date. We compared trends in case counts among detained persons, staff, and residents of Chicago during the study period by creating logarithmic-scale graphs of new and total cases; weekly averages were calculated to account for testing variation by day. All analyses were done using SAS v9.4 (Cary, North Carolina).

This study was reviewed by CDC, CDPH, CCH, and CCSO institutional review boards or the equivalent entity, and deemed not to be research involving human subjects and public health response.

## RESULTS

During March 1–April 30, 2020, 907 COVID-19 cases were identified among detained persons (n = 628, 69.2%) and staff (n = 279, 30.8%) (Figure 1). Of the 628 detained persons testing positive, 479 (76.3%) were symptomatic at time of specimen collection and 149 (23.7%) were identified through asymptomatic testing. All 279 staff cases reported symptoms. AR among detained persons was 12.9% (Table 1); median time at CCJ was 250 days (IQR 98□ 541), and 598 (95.2%) had been incarcerated or detained for >14 days at the time of their positive test.

**Table 1.**
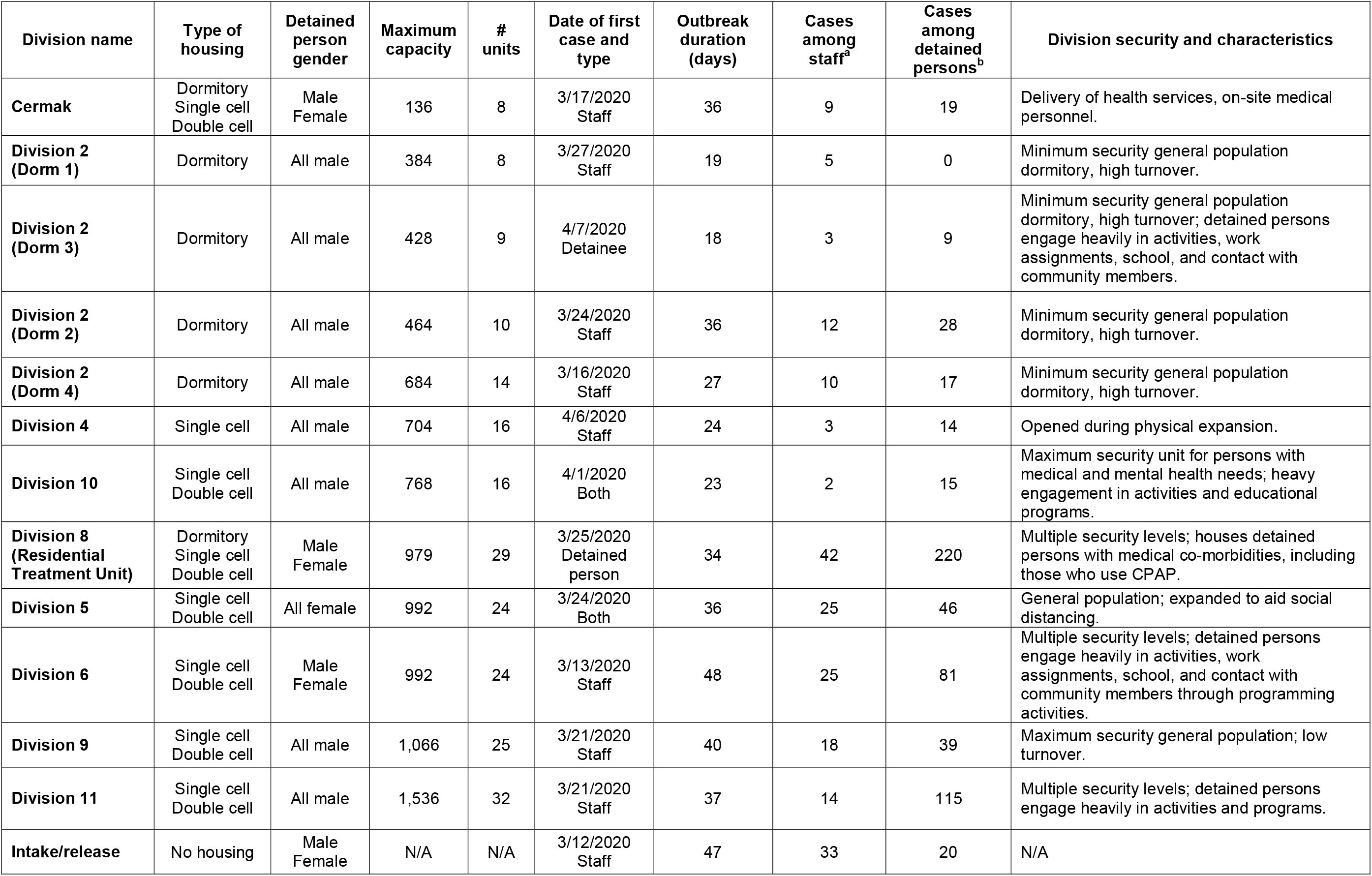
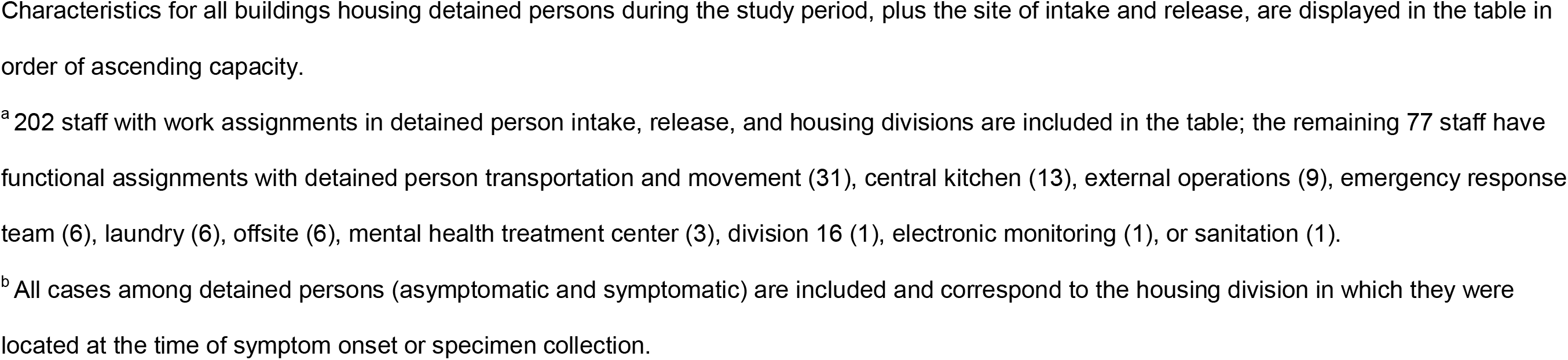
Housing characteristics and COVID cases in one of the largest jails in the United States by housing division—Cook County, IL, March 1– April 30, 2020

| Division name | Type of housing | Detained person gender | Maximum capacity | # units | Date of first case and type | Outbreak duration (days) | Cases among staff<sup>a</sup> | Cases among detained persons<sup>b</sup> | Division security and characteristics |
| --- | --- | --- | --- | --- | --- | --- | --- | --- | --- |
| **Cermak** | Dormitory<br>Single cell<br>Double cell | Male<br>Female | 136 | 8 | 3/17/2020<br>Staff | 36 | 9 | 19 | Delivery of health services, on-site medical personnel. |
| **Division 2 (Dorm 1)** | Dormitory | All male | 384 | 8 | 3/27/2020<br>Staff | 19 | 5 | 0 | Minimum security general population dormitory, high turnover. |
| **Division 2 (Dorm 3)** | Dormitory | All male | 428 | 9 | 4/7/2020<br>Detainee | 18 | 3 | 9 | Minimum security general population dormitory, high turnover; detained persons engage heavily in activities, work assignments, school, and contact with community members. |
| **Division 2 (Dorm 2)** | Dormitory | All male | 464 | 10 | 3/24/2020<br>Staff | 36 | 12 | 28 | Minimum security general population dormitory, high turnover. |
| **Division 2 (Dorm 4)** | Dormitory | All male | 684 | 14 | 3/16/2020<br>Staff | 27 | 10 | 17 | Minimum security general population dormitory, high turnover. |
| **Division 4** | Single cell | All male | 704 | 16 | 4/6/2020<br>Staff | 24 | 3 | 14 | Opened during physical expansion. |
| **Division 10** | Single cell<br>Double cell | All male | 768 | 16 | 4/1/2020<br>Both | 23 | 2 | 15 | Maximum security unit for persons with medical and mental health needs; heavy engagement in activities and educational programs. |
| **Division 8 (Residential Treatment Unit)** | Dormitory<br>Single cell<br>Double cell | Male<br>Female | 979 | 29 | 3/25/2020<br>Detained person | 34 | 42 | 220 | Multiple security levels; houses detained persons with medical co-morbidities, including those who use CPAP. |
| **Division 5** | Single cell<br>Double cell | All female | 992 | 24 | 3/24/2020<br>Both | 36 | 25 | 46 | General population; expanded to aid social distancing. |
| **Division 6** | Single cell<br>Double cell | Male<br>Female | 992 | 24 | 3/13/2020<br>Staff | 48 | 25 | 81 | Multiple security levels; detained persons engage heavily in activities, work assignments, school, and contact with community members through programming activities. |
| **Division 9** | Single cell<br>Double cell | All male | 1,066 | 25 | 3/21/2020<br>Staff | 40 | 18 | 39 | Maximum security general population; low turnover. |
| **Division 11** | Single cell<br>Double cell | All male | 1,536 | 32 | 3/21/2020<br>Staff | 37 | 14 | 115 | Multiple security levels; detained persons engage heavily in activities and programs. |
| **Intake/release** | No housing | Male<br>Female | N/A | N/A | 3/12/2020<br>Staff | 47 | 33 | 20 | N/A |

Characteristics for all buildings housing detained persons during the study period, plus the site of intake and release, are displayed in the table in order of ascending capacity.
<sup>a</sup> 202 staff with work assignments in detained person intake, release, and housing divisions are included in the table; the remaining 77 staff have functional assignments with detained person transportation and movement (31), central kitchen (13), external operations (9), emergency response team (6), laundry (6), offsite (6), mental health treatment center (3), division 16 (1), electronic monitoring (1), or sanitation (1).
<sup>b</sup> All cases among detained persons (asymptomatic and symptomatic) are included and correspond to the housing division in which they were located at the time of symptom onset or specimen collection.

**Figure 1.**
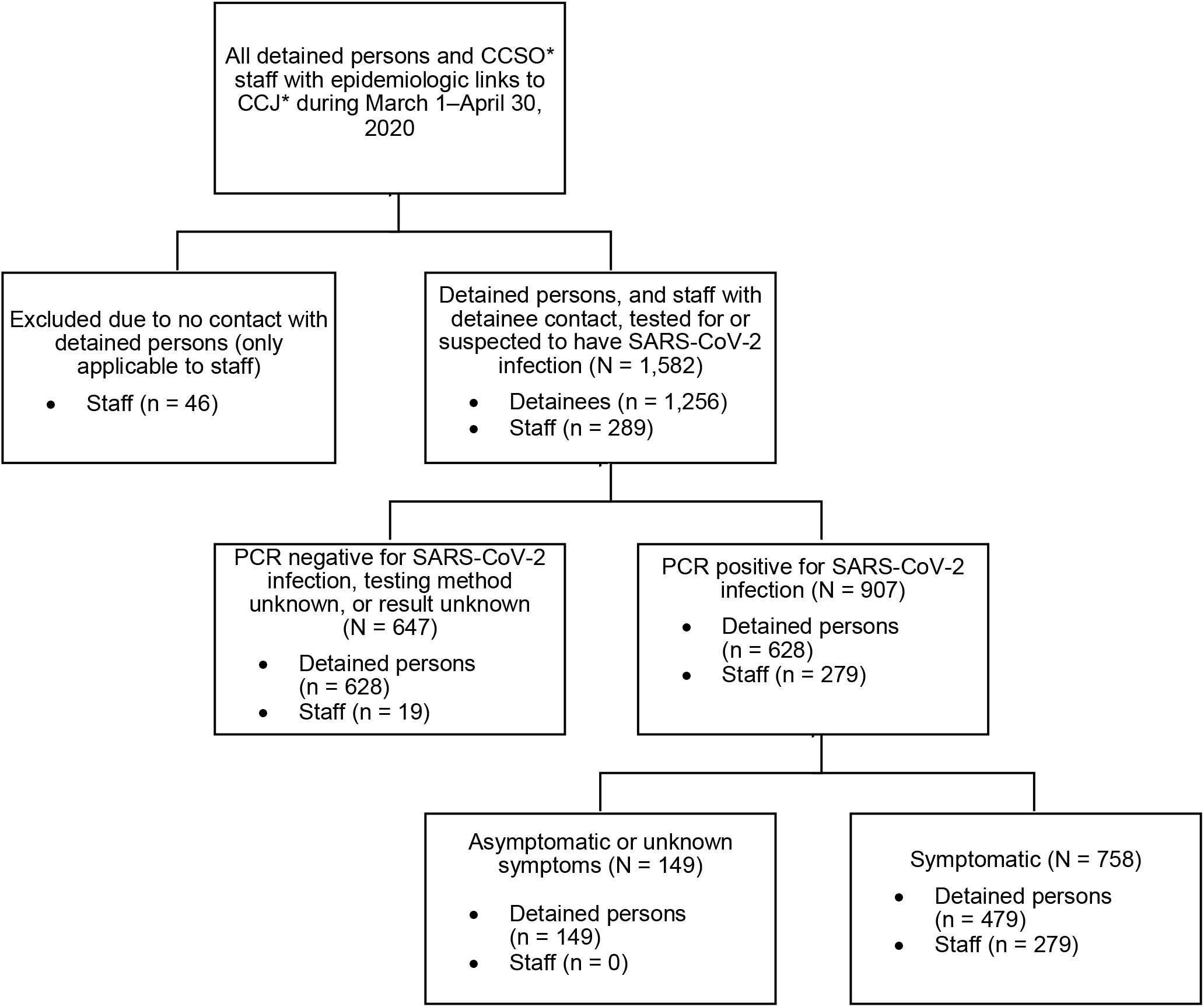
Criteria for inclusion in description of a COVID-19 outbreak in one of the largest jails in the United States—Cook County, IL, March 1–April 30, 2020. Abbreviations: CCSO (Cook County Sheriff’s Office); CCJ (Cook County Jail); PCR (polymerase chain reaction assay; SARS-CoV-2 (severe acute respiratory syndrome coronavirus 2). The average detainee census during the study period was 4,884; the average number of staff on site daily was approximately 1,500. Among 1,256 detained persons and 289 staff with detainee contact epidemiologically linked to CCJ, 479 symptomatic detained persons, 149 asymptomatic detained persons, and 279 symptomatic staff were PCR-positive for SARS-CoV-2 and included in analyses.

Symptomatic cases are included in epidemic curves (Figure 1) and asymptomatic cases are displayed by date and division (Supplemental Figure 1).

### SYMPTOMATIC CASES

Beginning January 21, screening of newly detained persons was expanded to include COVID-19 symptoms (cough, shortness of breath, fever) per CDC guidelines.^5^ The earliest reported date of symptom onset in a person later testing positive for SARS-CoV-2—a staff member in the transportation unit (Figure 2)—was March 2. In the week following identification of the first case in a detained person (March 18), 101 additional symptomatic cases (65 in detained persons, 36 in staff) were identified.

**Figure 2:**
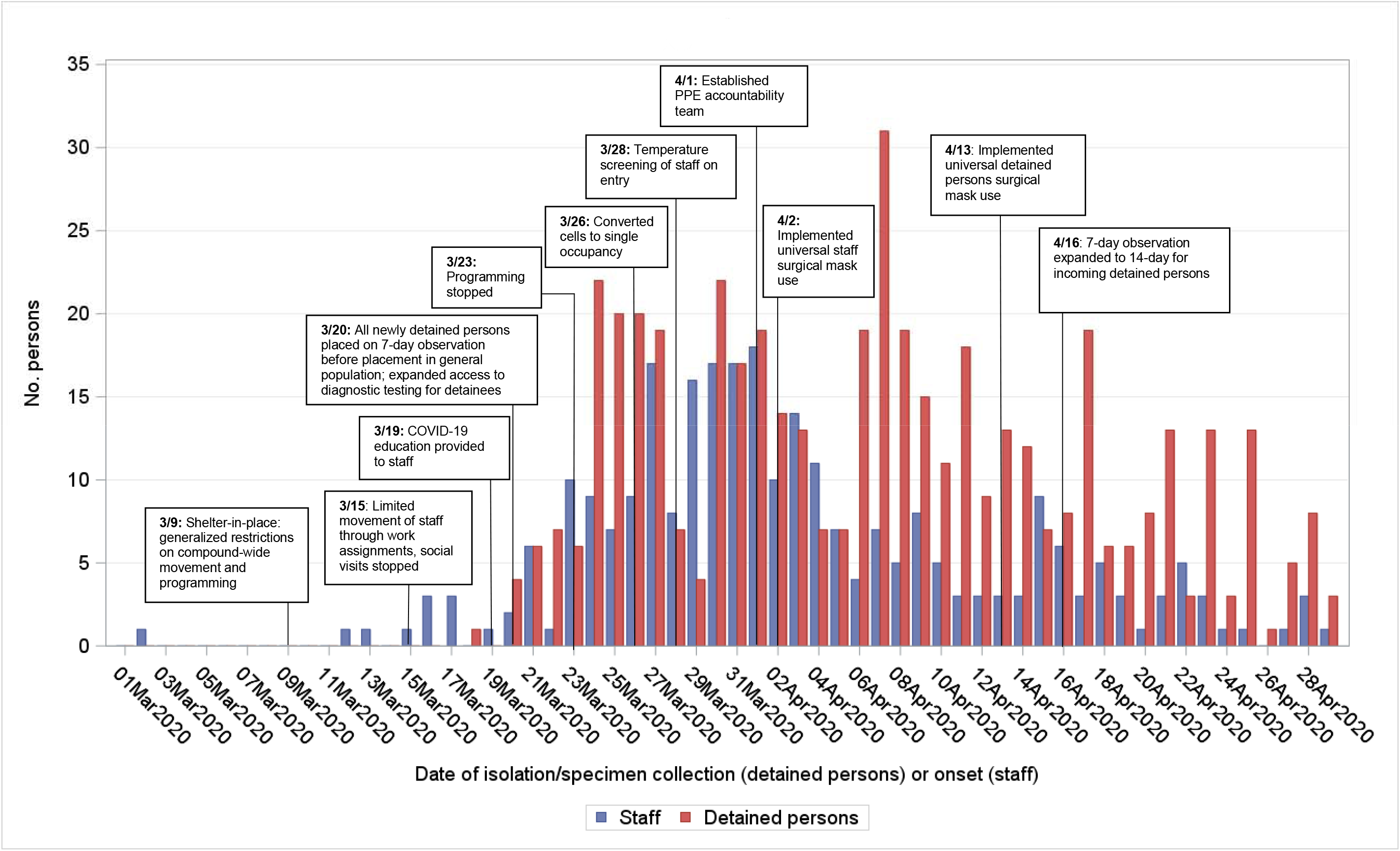
Number of symptomatic cases of COVID-19 by date of symptom onset among detained persons and staff with timeline of interventions in one of the largest jails in the United States—Cook County, IL, March 1–April 30, 2020 (n = 628) When constructing the epidemiologic curve, the date of medical isolation as a proxy for the date of symptom onset was used for detained persons, and self-reported symptom onset date was used for staff. Screening for influenza-like illness among incoming detained persons began October 1, 2019 and was expanded on January 21, 2020 to include symptoms of COVID-19 consistent with CDC guidelines. Screening of asymptomatic detained persons (not displayed in epidemic curve) began on March 3, 2020 among high-risk individuals in the Residential Treatment Unit; testing of all incoming detained persons upon intake began on April 20, 2020.

Symptom onset among all cases peaked March 30, with 39 symptomatic cases (22 in detained persons, 17 in staff). April 7 experienced the most symptomatic cases among detained persons (n = 31).

### INTERVENTIONS

Early interventions included enhanced cleaning and disinfection, eliminating aerosol-generating procedures (e.g., continuous positive airway pressure devices [CPAP]) in common areas (beginning March 20), hand hygiene education, and training staff on personal protective equipment (PPE) use.

CCJ began “sheltering in place” March 9, placing generalized restrictions on compound-wide movement and reducing programmatic activity. On March 15, social visitation ceased, and as of March 20, all newly detained persons were cohorted in small groups (10–30 individuals) under 7-day observation (extended to 14-day on April 16) to monitor symptoms before entering the general population. All programmatic activity was suspended on March 23.

CDPH conducted an on-site assessment on March 25 and provided guidance consistent with interventions already implemented by CHS/CCH. These included symptom screening (later supplemented by COVID-19 testing) at intake, immediate medical isolation of symptomatic individuals, social distancing, and quarantine of all detained persons on the unit for ≥14 days. Social distancing included spacing beds 6 feet apart in dormitories and reducing all cells to single occupancy beginning March 26 and completed by April 21. CDPH expanded infection control guidance for staff, including recommendations for PPE use based on task and cohorting by duty location (Figure 2).

Beginning March 28, all staff were screened for fever (99.4□ F/38□ C) and COVID-19 symptoms upon entry into CCJ.^5^ Presence of fever or symptoms required staff to abstain from work for 14 days. A PPE accountability team was assembled April 1, and staff were required to use surgical masks beginning April 2. Universal surgical mask use by detained persons during waking hours began April 13. On April 20, all newly detained persons were tested on intake using ID NOW in addition to undergoing 14-day quarantine (Figure 2).

### DIVISION CHARACTERISTICS AND EPIDEMIC CURVES

All nine housing divisions experienced cases despite variation in housing type, capacity, security, and characteristics (Table 1). Epidemic curves for certain divisions demonstrated a traditional bell-shape; others experienced sporadic cases (Figure 3). In 9/13 buildings, staff cases arose first, with a median 3 days between the first case in a staff member and a detained person.

**Figure 3:**
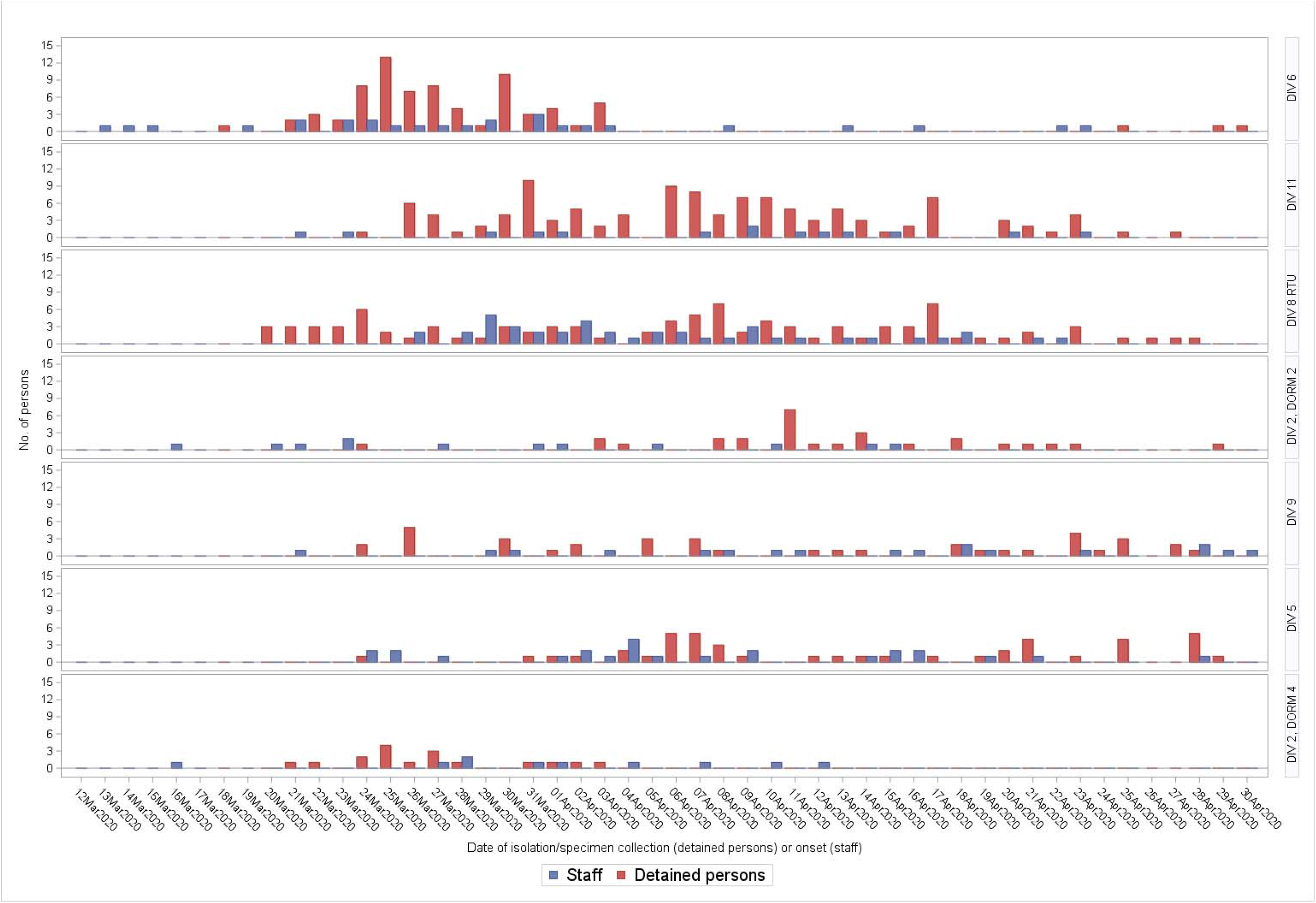
Number of symptomatic cases of COVID-19 by date of symptom onset among detained persons and staff by housing division in one of the largest jails in the United States—Cook County, IL, March 1–April 30, 2020. Epidemic curves for seven buildings representing six housing divisions (Division 6, Division 11, Division 8 [Residential Treatment Unit], Division 2 Dorm 2, Division 9, Division 5, and Division 2 Dorm 4) with COVID-19 cases among symptomatic staff and detainees are shown. Building names are labelled on the righthand side of each respective curve. Buildings are those with high case counts compared to remaining buildings not shown (Divisions 10 and 4, Division 2 Dorms 1 and 3, Cermak, and intake/release).

Division 6 had the highest level of programmatic activity and was the first to experience symptomatic cases among both groups. One hundred cases (75 among detained persons) were confirmed March 18– April 30 in Division 6. Detained persons were housed in double cells and had programming involving movement outside of their unit until March 19 (Figure 2).

The Residential Treatment Unit (RTU) housed more individuals with medical needs and at increased risk for COVID-19 medical complications than other divisions. RTU had the most symptomatic cases overall (137), including 42 among staff (15% of all staff cases) and 95 among symptomatic detained persons (20% of all symptomatic cases) (Figure 3).

### ASYMPTOMATIC CASES AMONG DETAINED PERSONS

In total, 631 asymptomatic detained persons were tested for SARS-CoV-2; 149 (23.6%) were positive, with percent positive ranging from 8% (2/25, Division 5) to 50% (125/249, RTU; Supplemental Figure 1). The unit with the highest percent positive was a dormitory with 37 individuals in the RTU, which housed individuals with comorbidities, including some who used CPAP until use in common areas was stopped. Of the 275 newly detained persons tested, 12 (4.8%) were positive. The 149 cases identified through asymptomatic testing represented 23.7% of all cases among detained persons at CCJ.

### FATALITIES

Seven detained persons and two staff died (case-fatality rate for both = 1.1%). Of fatal cases among detained persons, ages ranged from 42□ 64 years; all were male and had multiple comorbidities, most commonly hypertension, hyperlipidemia, and obesity.^10^

### OUTBREAK TRAJECTORY

Early in the outbreak, increases in cases among staff and detained persons paralleled that in Chicago, Illinois. After implementation of interventions, cases declined in detained persons and staff, even as cases increased dramatically in Chicago (Figure 4). Weekly averages demonstrated a decline in cases among detained persons a week after staff cases began declining.

**Figure 4:**
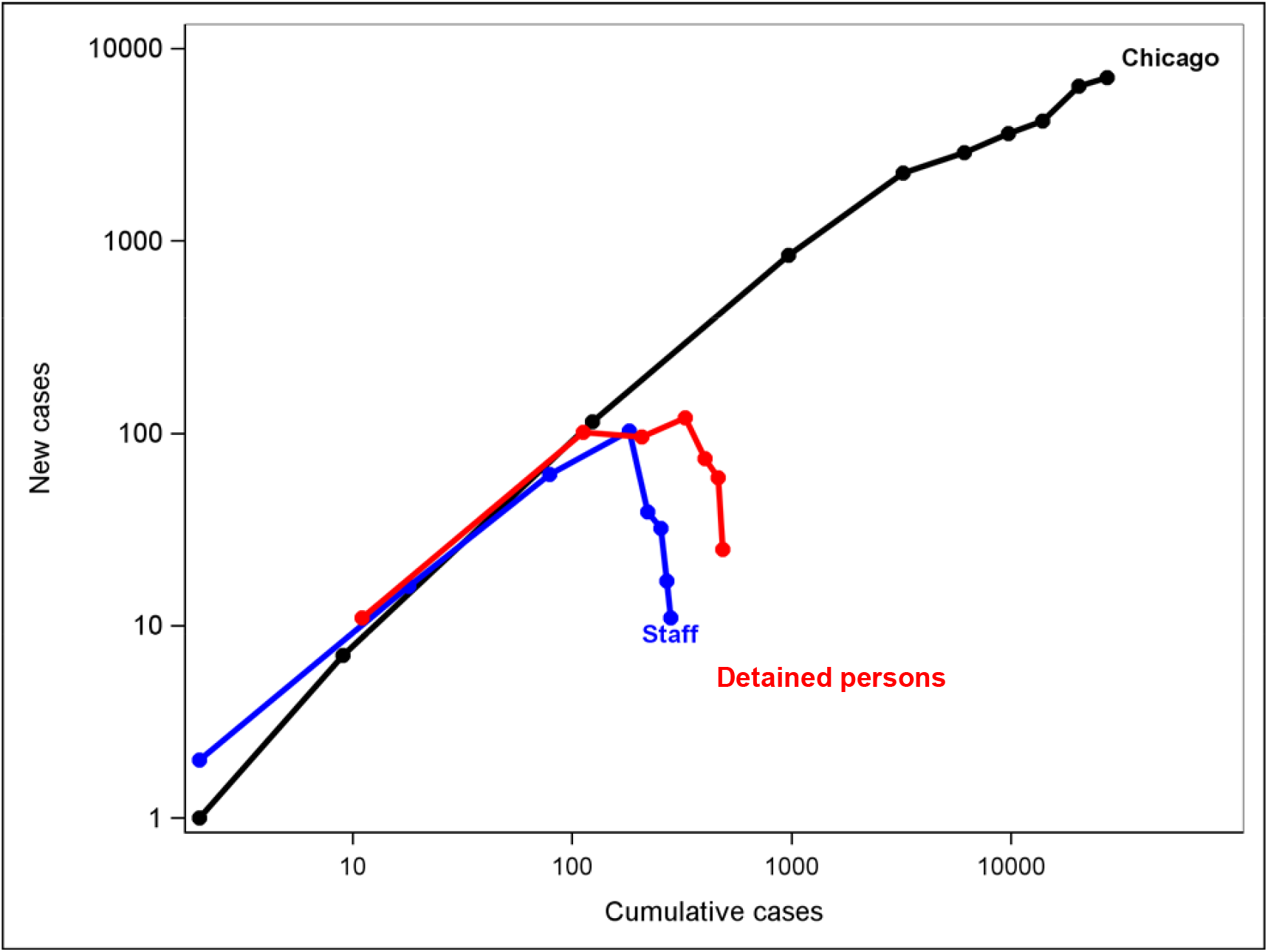
Ratio of new to cumulative cases among detainees and staff in Cook County Jail compared to Chicago—IL, March 1–April 30, 2020. Ratios of new cases to cumulative cases were calculated for each week of the study period among detained persons, staff, and residents of Chicago and plotted on a logarithmic scale to show outbreak trajectories. For staff, all symptomatic persons with validated molecular test results from the Illinois National Notifiable Disease Surveillance System (I-NEDSS) were included using date of onset as the referent time point; all asymptomatic and symptomatic detained persons testing PCR-positive were included using date of symptom onset or specimen collection as the referent time point. Data for all persons residing in the state of Illinois meeting the case definition for confirmed SARS-CoV-2 infection were extracted from the I-NEDSS system and included using the date of specimen collection. Each node represents 1 week of the study period; the highest number of total cases were identified in the jail the week of April 5^th^ and fell thereafter. The initial doubling times for Chicago, staff, and detained persons were 2.22, 2.15, and 2.1 days, respectively, represented by the increasing slope prior to peak for each population.

## DISCUSSION

Less than 2 months after the first COVID-19 case was identified in CCJ, almost 1,000 detained persons and staff had been infected with SARS-CoV-2. This represents an AR of nearly 13% among detained persons and occurred despite early adoption of containment and mitigation practices. This constitutes one of the largest outbreaks of COVID-19 in a congregate setting described to date, illustrating the difficulties of controlling spread in correctional and detention facilities. Estimates of influenza spread in enclosed populations have found similar ARs^9^ (13%); experience suggests viral respiratory pathogens like COVID-19 can cause sizeable epidemics in large jails despite implementation of public health interventions.^10^ Expanding CCJ’s footprint to facilitate physical distancing, limiting movement, and implementing expanded testing were complex and resource-intensive interventions, but effectively slowed spread relative to the surrounding community even as cases there surged. Implementing expanded diagnostic testing at key points, such as intake, helped limit new introductions of the virus.

Investigations into outbreaks of respiratory viruses in other correctional and detention facilities have identified visitors^11^ and persons transferred between facilities^12^ as possible sources. Restriction of movement within the jail was likely one of the most critical and timely interventions in gaining control of this outbreak; the division with the highest level of movement and most contact with individuals entering from the community experienced the earliest peak. Implementation and enforcement of social distancing of ≥6 feet, surgical mask use, increased access to soap and alcohol-based hand sanitizer, and enhanced cleaning and disinfection practices also likely reduce extent of spread. Later expansion of diagnostic testing, including at intake and of asymptomatic individuals, allowed for medical isolation of cases and reduction in spread. Enhanced measures including PPE accountability were likely also effective.

Our data suggest the important role that community-dwelling staff played in COVID-19 introductions into CCJ as cases among staff often preceded cases in detained persons. We also show the effectiveness of employee interventions despite inclusion of <100% of personnel. Implementation of universal screening for symptoms and temperature checks is important, but ensuring access to testing and compliance with illness reporting are vital, as are flexible and non-punitive leave policies to allow sick employees to stay home.

As with other outbreaks in correctional and detention facilities^13^, close cooperation between onsite medical service providers, law enforcement, and local and federal public health officials were critical to successful containment of COVID-19. Efforts to facilitate social distancing and medical isolation through expanding CCJ’s footprint likely reduced transmission.^14^ Physical distancing to the degree accomplished at CCJ may not be feasible in all facilities, but use of quarantine and cohort housing may be possible even in smaller, more restricted facilities.

A high proportion (23.6%) of exposed but asymptomatic detained persons were found to be positive, similar to other congregate settings such as homeless shelters.^15^ The role of these individuals in SARS-CoV-2 transmission is not well understood.^16^ Widespread testing facilitates rapid identification, early medical isolation, and reduction in potential for spread, though optimal timing for widespread testing is not known. Newly detained persons are exposed to the community prior to entering the jail, making expanded testing and cohorting at intake essential to limiting transmission.

This investigation has several limitations. First, testing capacity was limited early in the outbreak, potentially underestimating the number of cases; comprehensively employed mitigation methods reduced transmission even in the absence of full testing capacity. Our case definition required a positive PCR result; this may have excluded staff who were diagnosed clinically, or who had only serology performed. Further, while CCSO staff represented the largest group of staff members entering CCJ, other staff (e.g., healthcare staff) had a wide range of employers with no centralized listing and were not included in the study. Third, since demographic and clinical data were unavailable for cases other than those resulting in death, analyses by race, sex, and age could not be conducted. Lastly, because interventions were often implemented simultaneously, it was difficult to ascertain relative effectiveness.

## CONCLUSION

SARS-CoV-2 can spread rapidly in correctional and detention facilities, causing significant morbidity and mortality. Effective response to the COVID-19 outbreak at CCJ demonstrates the need for dynamic and aggressive application of intervention strategies, but also shows how timely response can reduce case counts and prevent morbidity and mortality in correctional or detention facilities.

## Data Availability

Data were provided by the Cook County Sheriff's Office, Chicago Department of Public Health, Cermak Health Services, and Cook County Health. Access to data submitted into the Illinois' National Electronic Disease Surveillance System was provided by Chicago Department of Public Health. Data represent protected health information (PHI), and cannot be made available in raw form. Results are presented in aggregate in this manuscript. Authors had access to data.

## ACKNOWLEDGMENTS

We would like to acknowledge the contributions of Cook County Sheriff Thomas J. Dart, Dr. Andrew Defuniak, Dr. Stamatia Richardson, Michael Miller, Brad Curry, Tarry Williams, and Linda Follenweider for their tireless efforts to implement these interventions. We would also like to acknowledge Jaqueline Tate, Kathryn Curran, Reena Doshi, Patrick Moonan, and the CDC field study team for their support. Mary Ann K. Hall, Sharon Saydah, and Louise K. Francoise Watkins contributed meaningfully to the review of this manuscript.

---

**Supplementary Figure 1.**
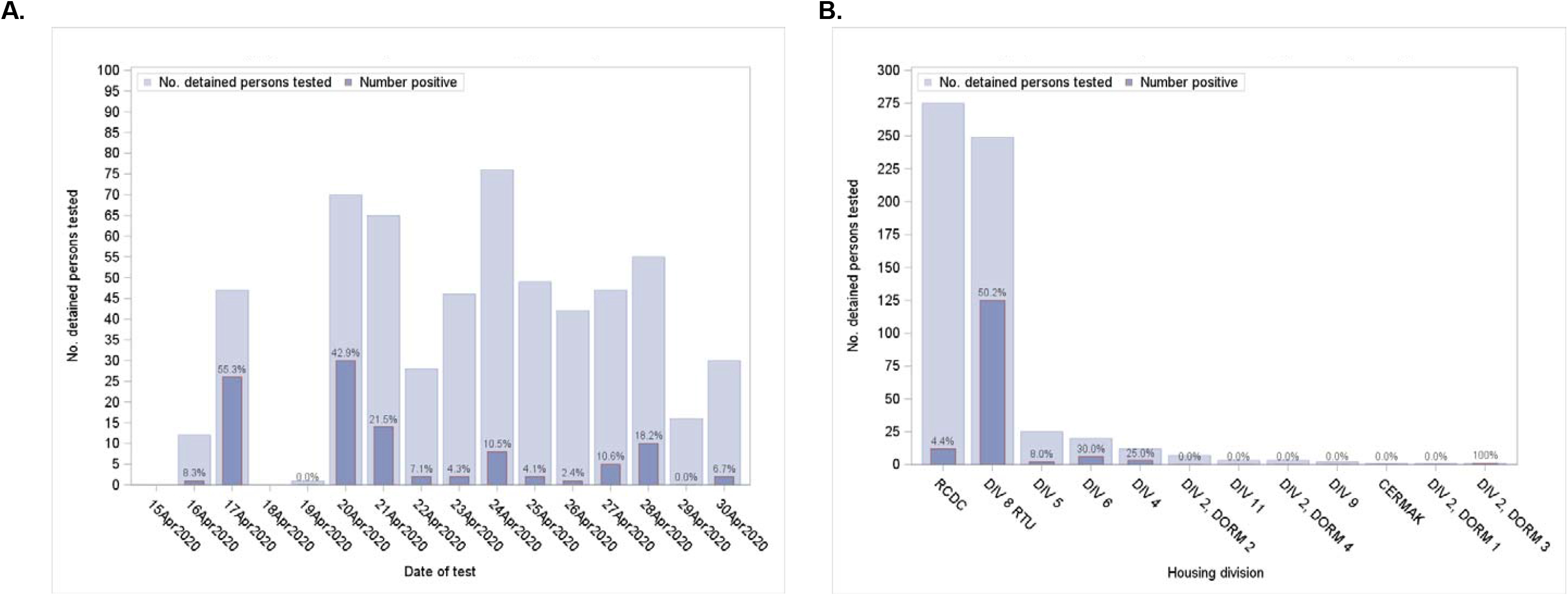
**(a)** Number of asymptomatic detained persons tested and percent positive by date of test*, and **(b)** number of asymptomatic detained persons tested and percent positive by housing division, in one of the largest jails in the United States—Cook County, IL, March 1–April 30, 2020. Light blue bars represent total number of asymptomatic detained persons tested for SARS-CoV-2 infection (a) on a given day or (b) while housed in a given building during the study period. Dark blue bars represent the total number positive, with data labels included to provide percent testing positive. *Data displayed in Figure 1A do not include asymptomatic detained persons housed in the Residential Treatment Unit (RTU) tested on March 25, 2020 (42 tested, 41 [97.6%] positive). Testing of asymptomatic detained persons began in earnest on April 16, 2020 and was expanded as capacity increased. RCDC denotes site of intake/release of detained persons.

